# The direct implementation costs of HIV pre-exposure prophylaxis in Lesotho and Zimbabwe: a costing study of PrEP choice involving oral pills, the dapivirine ring, and long-acting injectable cabotegravir to inform policy setting

**DOI:** 10.64898/2026.03.05.26347680

**Authors:** Joseph Corlis, Lori Bollinger, Collin Mangenah, Getrude Ncube, Nthuseng Marake-Raleie, Ramatsoai Soothoane, Emily Gwavava, Tatenda Yemeke, Marga Eichleay, Shubha Kapuganti, Peter Stegman, Nicole Bellows, Katharine Kripke

**Author notes:** Corresponding author (JC). Takoma Park, Maryland, United States of America. Glastonbury, Connecticut, United States of America. Durham, North Carolina, United States of America (Marga Eichleay). Rockville, Maryland, United States of America (Peter Stegman).

## Abstract

Because of its recent regulatory approval in southern and eastern Africa, CAB PrEP represents a scientific advancement with unknown implementation costs in most African countries. To our knowledge, this paper is the first study comparing PrEP costs in health facilities where clients had a choice between three PrEP methods. We collected and analyzed the direct service delivery costs for each method using the same costing approach and assumptions at three facilities in Lesotho and six facilities in Zimbabwe. On average, in Lesotho, the direct costs of providing CAB PrEP were $57.22 for an initiation visit and $54.20 for a refill visit (same PrEP product dose dispensed in both visit types), while the direct costs of oral PrEP were $22.47 (initiation visit with one month of PrEP dispensed) and $31.98 (refill visit dispensing a three-month dose of medication), and the direct costs of the dapivirine ring were $34.27 (initiation visit with one month of PrEP dispensed) and $50.70 (refill visit dispensing a three-month supply). In Zimbabwe, the average per-visit direct costs to provide CAB PrEP were $48.26 (initiation visit) and $47.40 (refill visit), to provide oral PrEP were $13.47 (initiation visit with one month of PrEP dispensed) and $21.78 (refill visit dispensing a three-month dose), and to provide the dapivirine ring were $42.56 (refill visit dispensing a three-month supply). Initiation visits for the dapivirine ring were not observed in Zimbabwe. At a time when national governments are creating budgets for the HIV response with decreased financial support from bilateral and multilateral partners, this paper will inform HIV prevention planning by providing critical client-level data from the healthcare provider perspective.

## Introduction

Most countries in sub-Saharan Africa, including Lesotho and Zimbabwe, have incorporated oral pre-exposure prophylaxis (PrEP) into their HIV prevention programs for nearly a decade [1, 2]. Oral PrEP, which requires users to take a daily pill, is highly effective at preventing HIV infections when used as directed; however, oral PrEP access, uptake, and adherence have been hindered by individual, familial, social, and structural barriers [1, 3–7]. Novel PrEP methods aim to address some of these barriers. The dapivirine ring—a once monthly method—is a discreet option for females that does not require daily dosing and has few side effects; however, its effect against HIV acquisition has been measured as 35-62% in clinical trials [8–10]. Another PrEP method, long-acting injectable cabotegravir (CAB PrEP), requires injections every 2 months. Evidence from the HPTN 083 and HPTN 084 trials demonstrated that CAB PrEP is superior to daily oral PrEP in preventing HIV acquisition among cisgender women in Africa, cisgender men who have sex with men, and transgender women [11, 12].

In Zimbabwe, tenofovir disoproxil fumarate-emtricitabine (TDF-FTC) is commonly prescribed for oral PrEP whereas in Lesotho tenofovir disoproxil fumarate/lamivudine (TDF/3TC) is typically given. In 2021, Zimbabwe became the first country globally to approve the dapivirine ring as a method of PrEP for HIV prevention and, the following year, it became the first country in Africa to grant regulatory approval of CAB PrEP [13]. Lesotho approved the dapivirine ring in 2023 and CAB PrEP in 2024 [14]. In both countries, the dapivirine ring and CAB PrEP have been provided through implementation studies and are not yet part of routine PrEP offered by the ministries of health.

International donor organizations such as the United States President’s Emergency Plan for AIDS Relief (PEPFAR) and the Global Fund to Fight AIDS, Tuberculosis and Malaria have historically co-financed the procurement and delivery of PrEP in many low- and middle-income countries, including Lesotho and Zimbabwe. Because primary costing studies are typically expensive to conduct and approvals for the dapivirine ring and CAB PrEP are recent, the direct service delivery costs of providing PrEP to clients in the context of routine implementation in settings where PrEP users are free to choose between multiple methods have not been accurately established in many countries [15]. Now that bilateral and multilateral donors are shifting the burden of funding and implementing HIV programs to domestic governments and their partners, the need for greater financial clarity is especially high [16]. As PrEP methods expand options for HIV prevention, understanding their service delivery costs is essential for informing policy and programmatic decisions.

## Methods

### Setting

Catalyzing Access to New Prevention Products to Stop HIV (CATALYST) was a large-scale study funded by PEPFAR through the United States Agency for International Development (USAID) that evaluated the implementation of PrEP choice among more than 6,000 women across 28 public health delivery sites [10]. Starting in 2024, the CATALYST study team planned to collect and analyze cost data related to oral PrEP, the daviripine ring, and CAB PrEP in three countries—Lesotho, Uganda, Zimbabwe. Most of the intended data were gathered in Lesotho and Zimbabwe before USAID issued a stop-work order in January 2025 that paused, and subsequently terminated, the study. With supplemental funding from an anonymous donor, we analyzed and present the data for Lesotho and Zimbabwe here.

### Cost data collection

This costing study took a healthcare provider perspective, focusing on direct service delivery financial costs (e.g., personnel time, equipment, consumables) for the provision of three PrEP methods. While the addition of dapivirine ring and CAB PrEP was novel in these sites and conducted as part of the CATALYST study, provision was per national guidelines and, therefore, simulated aniticipated routine provision. Between 9 September and 8 November 2024, data collectors used tablets to gather time-motion data for 166 clients receiving PrEP services across all three CATALYST-participating facilities in Lesotho and for 188 clients receiving PrEP services across all six CATALYST-participating facilities in Zimbabwe to estimate the resources used during service delivery. This convenience sampling of clients at the health facilities depended on client availability (i.e., facility throughput), facility staff identifying clients for potential participation, and data collector availability (i.e., not with other clients). The study aimed to collect time-motion data for at least five clients receiving each visit-type/method combination in each country. Collecting the minimum number of initiation and refill visits for the dapivirine ring was not possible in either country, in part, due to the low numbers of clients initiating ring once CAB PrEP was introduced.

After a obtaining written consent from participating clients, data collectors directly observed the clients during the entire health service visit, except for confidential parts of the consultation that could have caused the client discomfort. Observations indicated each step received in the national guidelines for PrEP service delivery (e.g., client registration, weight and vitals taken, group counseling, medical history taken, HIV testing including pre- and post-test counseling, PrEP eligibility screening, PrEP choice counseling, PrEP prescription given, PrEP dispensed, next appointment scheduled). For the steps in the service delivery process that the data collectors could not directly observe, health service providers recorded data on the steps provided, including start-stop times. These records were then handed to the data collectors. The data collectors also gathered salary data, commodity prices, laboratory costs, and normative expectations regarding all resources used in the different PrEP service delivery steps for each country.

### Data analysis

We conducted data analysis using Microsoft Excel to estimate a per-visit cost for each observed initiation and refill visit for each PrEP method for each country; we also estimated annual PrEP costs. We differentiated between visit types because the national guidelines are not the same for an initiation visit versus a refill visit (e.g., lab tests required, months of PrEP dispensed).

First, we compiled all 166 observations for Lesotho and 188 observations for Zimbabwe into the complete dataset. Next, we excluded seven observations from the dataset for Lesotho and 16 for Zimbabwe because either the visit type or PrEP method had missing data or multiple entries; this is discussed in more detail in the limitations section below. Then, we determined the duration of every step for each of the 331 observations (Lesotho n = 159, Zimbabwe n = 172) with complete data. For each cadre of personnel, annual salaries and benefits were converted to a per-minute cost, which we multiplied by the number of minutes each provider was observed participating in each client’s service visit. After accounting for any depreciation, equipment, furniture and reusable commodities were similarly converted to per-minute costs that were multiplied by the number of minutes for each step that indicated their use. The costs of single-use commodities were included in each step’s cost based on normative standards for the materials necessary to complete each observed step. To calculate annual costs for oral PrEP and the dapivirine ring, we assumed one initiation visit (dispensing a one-month supply of PrEP) and four refill visits (each dispensing a three-month supply of PrEP) in the year. For CAB PrEP, we assumed one initiation visit and six refill visits (each injecting the same dose of PrEP product) in the year [17]. Finally, we examined differences by visit type, PrEP method, and country. All currency values are presented in 2024 USD.

### Ethical considerations

Ethical approvals for this study were secured from the appropriate research review boards, including the Johns Hopkins Bloomberg School of Public Health (23517), the Lesotho National Health and Research Ethics Committee (ID 221-2022), the Medical Research Council of Zimbabwe (MRCZ/A/2996), and the Medicines Control Authority of Zimbabwe (MCAZ CT261/2023).

## Results

In Zimbabwe, the average direct costs for initiation and refill visits were lowest for oral PrEP at $13.47 and $21.78, respectively (Fig 1). While the costing team did not observe initiation visits for the dapivirine ring in Zimbabwe, as no new enrolling clients chose the ring during the data collection period, refill visits for the ring were $42.56. For CAB PrEP, initiation visits were $48.26 and refill visits were $47.40. In contrast, in Lesotho, the average direct costs were higher: oral PrEP was $22.47 for initiation visits and $31.98 for refill visits, followed by the dapivirine ring at $34.27 for initiation visits and $50.70 for refill visits, and then CAB PrEP at $57.22 for initiation visits and $54.20 for refill visits.

**Fig 1.**
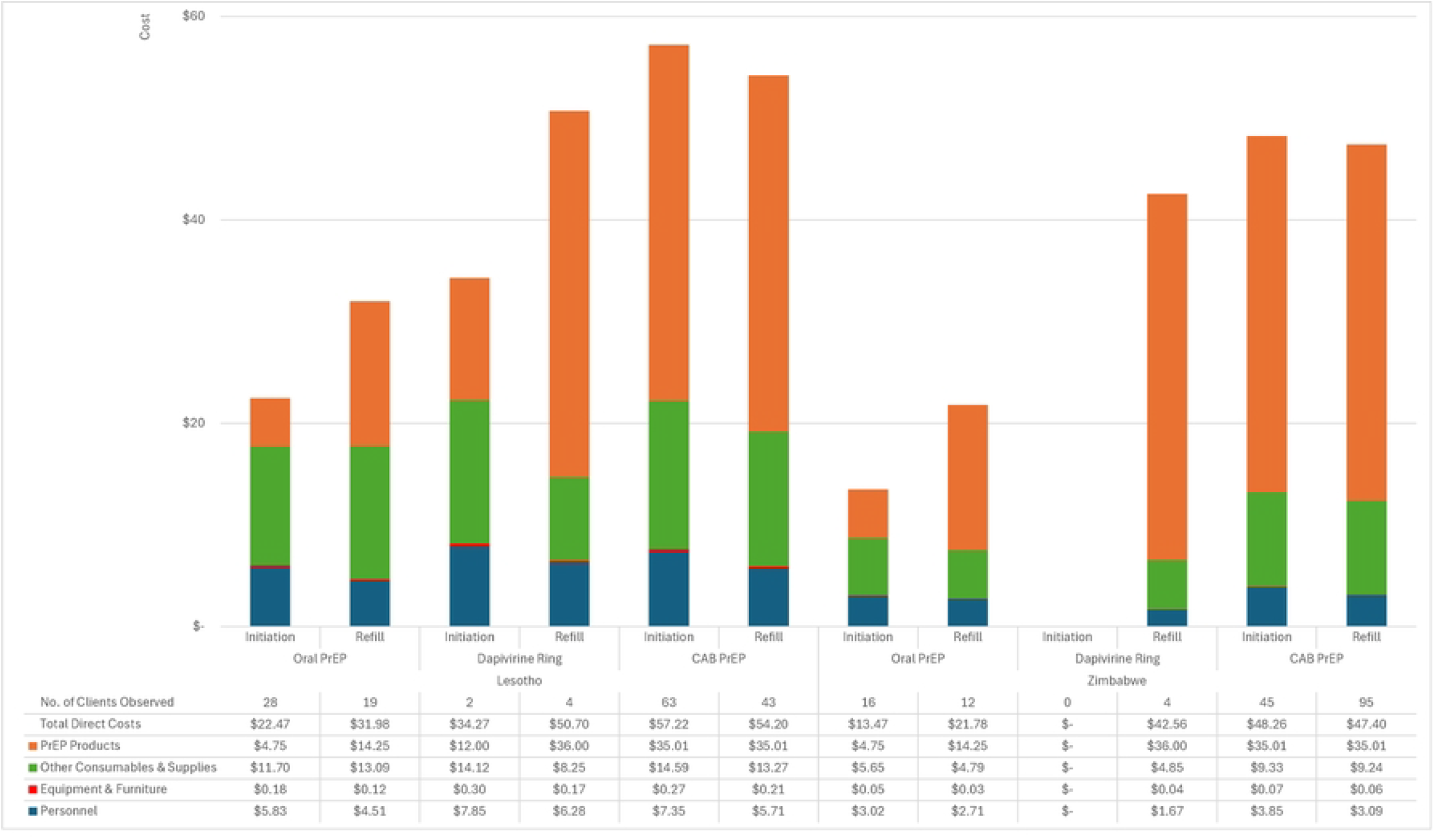
Average direct costs for PrEP provision in Lesotho and Zimbabwe by PrEP method, visit type, and cost category (2024 USD).

Setting aside the costs of the PrEP products, direct costs were higher in both countries for initiation visits than refill visits for all PrEP methods except oral PrEP in Lesotho, which was $0.01 more expensive on average. In Zimbabwe, the largest cost driver was PrEP products (35-85% of all direct costs) followed by other consumables and supplies (11-42% of all direct costs). In Lesotho, the largest cost driver was PrEP products for all CAB PrEP visits and for refill visits for oral PrEP and the dapivirine ring (45-71% of all direct costs). For initiation visits for oral PrEP and the dapivirine ring, the largest cost driver in Lesotho was other consumables and supplies (e.g., HIV tests, vacutainers, gloves, forms, pregnancy tests), which comprised between 41-52% of all direct costs.

Each PrEP method confers a different duration of protection against HIV infection. We estimated that average normative annual direct costs ranged between $150-$382 in Lesotho and $101-$333 in Zimbabwe (Table 1). We note that, because we did not observe initiation visits for the dapivirine ring in Zimbabwe, annual costs could not be calculated. In both countries, our estimates show CAB PrEP to be the most expensive PrEP method annually because, if clients attend all scheduled PrEP visits in accordance with national guidelines (i.e., do not need to restart), CAB PrEP requires more total visits in a year than oral PrEP or the dapivirine ring.

**Table 1:** Estimated average annual direct costs for each PrEP method with min-max range (2024 USD).

|  | **Lesotho (min - max)** | **Zimbabwe (min - max)** |
| --- | --- | --- |
| **Oral PrEP** | \$150.39 (\$117.87 - \$226.15) | \$100.59 (\$84.71 - \$120.70) |
| **Dapivirine Ring** | \$237.07 (\$220.59 - \$251.86) | - |
| **CAB PrEP** | \$382.41 (\$326.21 - \$498.50) | \$332.66 (\$110.80 - \$607.50) |

## Discussion

These estimates align with previous findings of oral PrEP costs in sub-Saharan Africa [18–20]. In Zambia, where the total costs for oral PrEP programming were calculated including demand creation and client support activities, direct service delivery costs represented 35-65% of the total costs for PrEP programming [21]. If this proportion is similar in Lesotho or Zimbabwe, then total oral PrEP service delivery costs likely exceed $175 per user per year.

While our annual cost estimates assume continuous PrEP use throughout the year, real-world utilization patterns show high levels of discontinuation of PrEP among women throughout the year [22–24]. When we applied the continuation rates reported by Ogalla et al. [22] to our oral PrEP visit costs, the average estimated annual cost per person initiated was $43 in Lesotho and $29 in Zimbabwe, not accounting for the possibility of reinitiating during the year. These figures represent approximately 30% of the normative annual costs for oral PrEP of $150 and $101, respectively. This distinction is critical for budget planning and resource allocation, as healthcare systems must account for both the costs of initiating individuals on PrEP and the reality that most will discontinue treatment during the year, although some may reinitiate PrEP later. Further analyses of the patterns of PrEP use within CATALYST and other studies could better inform estimates of the actual cost per person initiated or per person accessing PrEP per year.

As health policymakers and program planners face increasing scarcity of financial resources, PrEP may not be as affordable or cost-effective as other investments in health [25]. However, as the options for PrEP methods grow and people are able to choose methods that align with their preferences, the cost-effectiveness of the intervention may continue to improve. Lenacapavir, for example, is a twice annual injection that confers highly effective protection against HIV acquisition, which recently began gaining approval by country governments as a method for HIV prevention and treatment [26, 27]. The annual product cost of Lenacapavir is expected to be close to that of generic oral PrEP [28].

We note that this study has several limitations. First, service providers recorded some of the data for this study because data collectors were not permitted to observe clients in private consultation rooms; therefore, it is possible that data entry errors occurred that our study’s data collectors could not identify or remedy. When PrEP method or visit type was unclear (4.2% of observed clients in Lesotho, 8.5% in Zimbabwe), we censored observations from our dataset. Similarly, some of the steps conducted and recorded during the client service visits by providers did not relate to PrEP (e.g., family planning method prescription and dispensing); we did not include costs for those steps in the individual or average PrEP direct service costs. Additionally, because the time-motion study followed clients rather than providers, data collectors did not observe the non-client-facing time spent by providers to complete the PrEP service visit without the client present (e.g., charting, reviewing lab results). We could not generate an *ex post* calculation of non-client facing provider time because no data categorizing provider activities as client-facing versus non-client facing were collected as part of the study [29]. Without non-client-facing provider time, this study underestimates personnel costs in the calculations of each PrEP method. Finally, although the study initially intended to include client observations in Uganda and indirect costs for PrEP services in all countries, the data collectors were not able to collect that information before the study was terminated.

## Conclusions

This study is important because to our knowledge it is the first instance of costing PrEP choice in routine implementation. In Lesotho and Zimbabwe, women seeking to avoid HIV infection by using PrEP have a growing array of methods available to meet their needs based on preference and affordability. The direct costs borne by health facilities during PrEP initiation and refill visits are significant, but PrEP serves a critical role in country HIV prevention programs. We recommend that future PrEP studies report costs disaggregated by initiation and refill visits, take continuation and restart rates into account in the analysis, and include the non-client-facing provider time in estimates of facility personnel participation in PrEP service delivery.

## Data Availability

This study used third-party data. They were collected as part of the CATALYST study and, in accordance with the study’s research protocol, belong solely to the ministries of health of Lesotho and Zimbabwe. To request access to the data, readers can contact Dr. Nthuseng Marake at Lesotho’s Ministry of Health and Social Welfare and Dr. Owen Mugurungi at Zimbabwe’s Ministry of Health and Child Care.

## Acknowledgements

The authors thank Dr. Steven Forsythe for his contributions to the conceptualization of the costing study and its description in the CATALYST protocol.

## Author contributions

Conceptualization: Stegman P, Kripke K.

Data curation: Corlis J, Bollinger L, Kapuganti S, Stegman P, Bellows N.

Formal analysis: Corlis J, Bollinger L, Kapuganti S, Stegman P, Bellows N.

Funding acquisition: Eichleay M, Kripke K.

Investigation: Mangenah C, Soothoane R, Gwavava E, Yemeke T, Stegman P.

Methodology: Bollinger L, Yemeke T, Eichleay M, Stegman P, Bellows N, Kripke K.

Project administration: Gwavava E, Eichleay M, Stegman P, Bellows N, Kripke K.

Resources: Ncube G, Marake-Raleie N, Yemeke T, Eichleay M.

Supervision: Eichleay M, Stegman P, Kripke K. Validation: Bollinger L.

Visualization: Corlis J.

Writing – original draft: Corlis J.

Writing – review and editing: Bollinger L, Mangenah C, Ncube G, Marake-Raleie N, Soothoane R, Gwavava E, Yemeke T, Eichleay M, Kapuganti S, Stegman P, Bellows N, Kripke K.

